# Diagnosing hyperglycemia (GDM) in pregnancy: closing the door after the horse has bolted?

**DOI:** 10.1101/2024.10.21.24315904

**Authors:** Chittaranjan S. Yajnik, Souvik Bandyopadhyay, Dattatray S. Bhat, Rucha S. H. Wagh, Pallavi C. Yajnik, Rasika Ladkat, Kurus Coyaji, Caroline H. D. Fall

## Abstract

**Introduction:** A common belief is that gestational hyperglycemia (‘GDM’) develops during pregnancy and remits after delivery. It increases the risk of diabetes and obesity in the offspring and fuels the intergenerational epidemic of diabesity in the young. Intensive glycemic treatment in pregnancy RCTs failed to prevent this transmission because the critical pre- and peri-conceptional window for epigenetic programming was missed. Some studies have reported that women diagnosed with GDM were already hyperglycemic, obese, and insulin resistant before pregnancy. However, little is known about the life-course evolution of glycemia before the diagnosis of pregnancy hyperglycemia. The Pune Maternal Nutrition Study (PMNS) birth cohort has measured plasma glucose serially throughout childhood, puberty, young adulthood, pregnancy, and later, providing a unique opportunity to test the hypothesis that pregnancy hyperglycemia is only a window in lifetime hyperglycemia.

**Methods:** The PMNS birth cohort, established in 1993, included serial glucose measurements at ages 6, 12, and 18 years, as well as for females during pregnancy and post-delivery follow-up. Of 366 female cohort members, 171 became pregnant and delivered by February 2020. Given the small number of GDM (IADPSG criteria) we defined pregnancy hyperglycemia as the upper quartile (Q4) of fasting plasma glucose (FPG) and area under the curve (AUC) during an OGTT at 28 weeks gestation.

**Results:** At 28wks gestation these women were young (mean age 20.9y) and had a median BMI 21.7 kg/m^2^ [IQR, 20.0, 23.8]; 44 women had fasting hyperglycemia (FPG >4.7 mmol/l) and 39 had AUC hyperglycemia (AUC > 8.57 x 10^2^). For both groups, hyperglycemic women had higher glycemia from childhood through post-delivery compared to normoglycemic women, and higher HbA1c before pregnancy. Having an FPG above the highest quartile from childhood increased the odds of pregnancy hyperglycemia 2.22 times (95% CI 1.45, 3.38), and post-delivery glucose intolerance 5.22 times (2.40, 11.33); for AUC, the odds were 2.88 (1.31, 6.28) and 3.50 (1.36, 8.97) respectively.

**Interpretation:** Pregnancy hyperglycemia reflects persistent hyperglycemia since childhood. Diagnosing and managing hyperglycemia (‘GDM’) in pregnancy does not prevent exposure of the ovum and early embryo to an abnormal metabolic milieu and will not curtail the escalating epidemic of diabesity in the offspring. A pre-conceptional ‘primordial’ approach is essential.

**Research in context:** *Evidence before this study:*

- Most clinicians and researchers believe that ‘gestational’ diabetes develops during pregnancy and remits after delivery
- Some studies have reported higher glucose, HbA1c, lipids, and BMI years before diagnosis of GDM but these are underplayed as ‘risk factors’
- Randomised controlled trials of intensive glycemic control in pregnancy (usually initiated in the third trimester) do not prevent the long-term risk of diabetes and obesity in the offspring
- This may be partly due to genetic transmission but more likely due to pre- and peri-conceptional epigenetic programming due to maternal metabolic status

*What’s new in this study:*

- We describe for the first time a life course trajectory of glycemia and body size in women with pregnancy hyperglycemia, in the Pune Maternal Nutrition Study, a preconceptional birth cohort initiated 30 years ago
- Women with pregnancy hyperglycemia had consistently elevated glycemia from childhood, puberty, and young adulthood through pregnancy and post-pregnancy
- This demonstrates that pregnancy hyperglycemia is only a window in the life course hyperglycemia and not a *de novo* phenomenon.

*Implications of all the available evidence:*

- Our findings suggest that primordial prevention of the intergenerational vicious cycle of diabetes and obesity may be achieved by management of metabolic abnormalities before pregnancy
- This will shift the focus from the clinic to the community, from clinical medicine to public health
- Further research will define the role of genetic and epigenetic factors involved

## Introduction

The IDF Atlas 2025 estimates that every year 23 million babies (19.7 % of all pregnancies) in the world are born in a ‘hyperglycemic’ pregnancy, and an overwhelming majority of these are first discovered only during pregnancy, so-called gestational diabetes (GDM) (1). The majority of GDM cases are in low-and middle-income countries (LMICs). Current standards of practice recommend diagnosing GDM at 24-28 weeks of gestation (2). In addition to adverse pregnancy outcomes involving both the mother and the baby (3), GDM increases risk of diabetes, obesity and cardiovascular disease (CVD) in the child (4–6), referred to as the ‘vicious cycle of diabetes’ (7). RCTs of intensive treatment of GDM have shown a reduction in the incidence of large for gestational-age (LGA) / macrosomia and associated obstetric problems but have failed to protect the offspring against childhood obesity and glucose intolerance (8–11). This could be ascribed to genetic transmission and/or to fetal epigenetic programming of these conditions (12). Pima Indian data favoured the latter over genetic factors (6)

GDM is generally viewed as a transitory phenomenon, due to the metabolic-endocrine stress of pregnancy (4). It is a general belief that it develops during pregnancy and resolves after the delivery of the child. There is increasing evidence that the metabolic abnormalities of GDM (hyperglycemia, low insulin secretion, and insulin resistance) precede pregnancy. This was reported in the pioneering serial metabolic studies before and during pregnancy by Catalano (13), in clinic-based data in California (USA) (14), in the CARDIA (USA) (15) and MUSP (Australia) cohorts (16), in the Cardiovascular Risk in Young Finns Study (17), and a population-based study in Ontario Canada (18). However, we could not find any reports in the literature of a life course trajectory of glycemic measurements in women diagnosed with pregnancy hyperglycemia.

The Pune Maternal Nutrition Study (PMNS) is a pre-conceptional birth cohort study in six villages near Pune, India, with a 30-year follow-up since its inception in 1993 (19, 20). Data are available on 3 successive generations (F0, F1, F2). In the female offspring (F1) born in the PMNS, serial measurements of body size and glucose-insulin metabolism are available from birth, through childhood, puberty and adolescence, to their first pregnancy and after delivery. Glucose tolerance information is also available in their mothers (F0) when pregnant with the F1 child. This provided us with a unique opportunity to construct a life course trajectory of glycemia in F1 mothers with pregnancy hyperglycemia. We tested the hypothesis that pregnancy hyperglycemia originates in early life, not during pregnancy. This may help assess the appropriateness of the current clinical practice of diagnosing and treating GDM in pregnancy, and its implications for intergenerational transfer of risk of diabesity and the potential window for its primordial prevention.

## Methods

The PMNS was established in 1993 in six villages near Pune, India to study determinants of fetal growth and to study life-course evolution of cardio-metabolic risk factors (Figure 1) (19).

**Figure 1:**
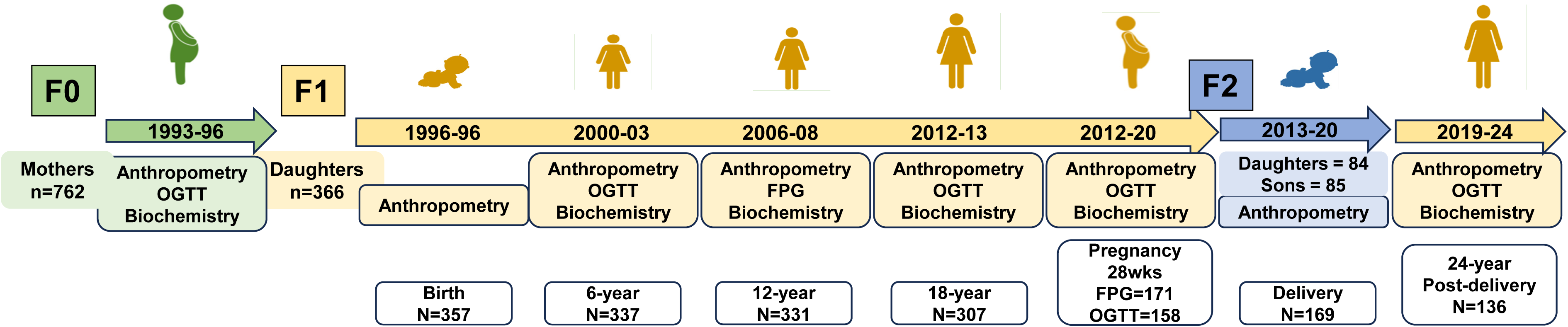
The Pune Maternal Nutrition Study. F0 mothers were studied before and during pregnancy, and the F1 children were born. F1 daughters were serially followed up from birth to 24-years of age including during pregnancy and post-pregnancy, with anthropometry and cardiometabolic measurements. Their children form the F2 generation. This paper traces glucose metabolism in F1 daughters from their intrauterine life, through childhood, puberty, adulthood and into pregnancy and post-pregnancy.

### F0 generation

Briefly, married, non-pregnant women (N=2,466) were followed up regularly for menstrual history and body size measurements (ESM fig. 1). They were recruited into the main study when a singleton pregnancy of <21 weeks’ gestation was confirmed by ultrasound. Maternal medical, nutritional, and biochemical information was obtained at 18- and 28-weeks’ gestation, including a 75gm fasting OGTT at 28-weeks. Deliveries happened at home or local health centres.

### Lifecourse measurements of F1 generation from birth to 18-years

Anthropometric measurements were made using standardized protocols at birth and during subsequent follow-ups (ESM table 1) (20). At age 6 years, a 1.75g/kg oral glucose tolerance test (OGTT), and at 18 years a 75g OGTT were performed (fasting, 30- and 120-min venous blood samples). At 12 years, only a fasting blood sample was collected. Plasma glucose was measured by GOD-POD method on an automated system, within and between batch CV was <3% (ESM table 2). HbA1c was measured on Bio-Rad D10 system (Bio-Rad labs, Hercules, CA, USA) between 15- and 17-years of age.

**Table 1:** Comparison of biomarkers between F1 participants with normoglycemia and hyperglycemia in pregnancy (based on quartiles of FPG and OGTT AUC at 28 weeks of gestation)

|  | Fasting plasma glucose |  |  | OGTT-AUC |  |  |
| --- | --- | --- | --- | --- | --- | --- |
|  | Normoglycemia | Hyperglycemia | Mean Z difference<br>(95% CI) | Normoglycemia | Hyperglycemia | Mean Z difference<br>(95% CI) |
| ***F0 generation (Mother)*** | *n*=108 | *n*=35 |  | *n*=98 | *n*=27 |  |
| FPG at 28wk gestation (mmol/l) | 3.9 | 3.9 | 0.08 (-0.32, 0.47) | 3.9 | 3.9 | -0.02 (-0.47, 0.44) |
| 2h glucose at 28wk gestation<br>(mmol/l) | 4.2 | 4.3 | -0.09 (-0.58, 0.39) | 4.2 | 4.5 | 0.23 (-0.22, 0.68) |
| ***F1 generation*** |  |  |  |  |  |  |
| ***Birth*** | *n*=125 | *n*=44 |  | *n*=113 | *n*=34 |  |
| Gestation (weeks) | 39.1 | 38.7 | -0.08 (-0.63, 0.46) | 39 | 38.5 | -0.2 (-0.71, 0.31) |
| Weight (g) | 2600 | 2500 | -0.07 (-0.43, 0.29) | 2550 | 2475 | -0.42 (-0.78, -0.05) |
| Length (cm) | 47.4 | 47.7 | -0.01 (-0.36, 0.34) | 47.4 | 46.8 | -0.43 (-0.82, -0.03) |
| ***2 Years*** | *n*=115 | *n*=38 |  | *n*=108 | *n*=32 |  |
| Height (cm) | **80.7** | **81.8** | **0.41 (0.05, 0.79)** | 81.2 | 80.1 | -0.27 (-0.69, 0.16) |
| Weight (kg) | **9.1** | **9.7** | **0.43 (0.06, 0.79)** | 9.2 | 9.1 | -0.17 (-0.58, 0.23) |
| ***6 Years*** | *n*=126 | *n*=44 |  | *n*=119 | *n*=38 |  |
| Height (cm) | **108.8** | **110.6** | **0.34 (0.00, 0.67)** | 109.5 | 108.3 | -0.17 (-0.56, 0.23) |
| Weight (kg) | **15.3** | **16.5** | **0.44 (0.10, 0.78)** | 15.6 | 15.5 | -0.04 (-0.44, 0.35) |
| BMI (kg/m<sup>2</sup>) | 12.9 | 13.4 | 0.32 (-0.03, 0.67) | 13 | 12.9 | 0.11 (-0.29, 0.51) |
| Waist circumference (cm) | **49.5** | **50.4** | **0.42 (0.07, 0.76)** | 50 | 49.8 | -0.05 (-0.46, 0.36) |
| Fasting glucose (mmol/l) | **4.7** | **5.1** | **0.42 (0.08, 0.77)** | 4.8 | 4.9 | 0.03 (-0.36, 0.41) |
| 30-min glucose (mmol/l) | 8.3 | 8.5 | 0.06 (-0.30, 0.42) | 8.2 | 8.6 | 0.23 (-0.15, 0.62) |
| 120-min glucose (mmol/l) | 5.6 | 5.7 | 0.23 (-0.11, 0.58) | 5.7 | 5.7 | 0.29 (-0.03, 0.61) |
| OGTT AUC | 8.31 | 8.55 | 0.16 (-0.19, 0.51) | 8.27 | 8.69 | 0.31 (-0.04, 0.66) |
| **12 Years** | *n* =125 | *n* =44 |  | *n* =118 | *n* =38 |  |
| Height (cm) | 139.3 | 142.1 | 0.21 (-0.12, 0.55) | 139.3 | 140.7 | 0.05 (-0.31, 0.42) |
| Weight (kg) | **27.1** | **29.4** | **0.36 (0.10, 0.78)** | 27.8 | 28.9 | 0.09 (-0.29, 0.47) |
| BMI (kg/m<sup>2</sup>) | **14.2** | **14.7** | **0.42 (0.07, 0.77)** | 14.4 | 14.3 | 0.01 (-0.41, 0.43) |
| Waist circumference (cm) | **55.5** | **56.3** | **0.35 (0.00, 0.70)** | 56 | 55.6 | 0.05 (-0.36, 0.45) |
| Fasting glucose (mmol/l) | **4.6** | **4.8** | **0.38 (0.04, 0.71)** | 4.7 | 4.7 | 0.04 (-0.30, 0.38) |
|  | *n* =120 | *n* =43 |  |  |  |  |
| Pre-pregnancy HbA1c (%),<br>*n*=163 | **5.3** | **5.5** | **0.27 (-0.07, 0.62)** | **5.3** | **5.5** | **0.11 (-0.02, 0.25)** |
| Pre-pregnancy HbA1c<br>(mmol/mol), *n*=163 | **34** | **37** | **0.27 (-0.07, 0.62)** | **34** | **37** | **0.11 (-0.02, 0.25)** |
| **18 Years** | *n* =120 | *n* =42 |  | *n* =113 | *n* =37 |  |
| Height (cm) | 156.8 | 158.5 | 0.20 (-0.15, 0.54) | 157.4 | 156.2 | -0.39 (-0.81, 0.03) |
| Weight (kg) | **43.9** | **47.3** | **0.50 (0.15, 0.85)** | 45.6 | 43.9 | -0.25 (-0.65, 0.15) |
| BMI (kg/m<sup>2</sup>) | **17.9** | **19.1** | **0.46 (0.11, 0.81)** | 18 | 18.3 | -0.05 (-0.50, 0.39) |
| Underweight | **76 (63.3)** | **15 (35.7)** | ***p*<0.05** | 63 (55.8) | 20 (54.1) | ns |
| Overweight/obese | **4 (3.3)** | **3 (7.1)** | ***p*<0.05** | 5 (4.4) | 2 (5.4) | ns |
| Waist circumference (cm) | **66.1** | **68.8** | **0.47 (0.13, 0.82)** | 67.5 | 66.9 | -0.18 (-0.57, 0.21) |
| Central obesity, ≥80 N(%) | 4 (3.3) | 3 (7.1) | ns | 6 (5.3) | 1 (2.7) | ns |
| Fasting glucose (mmol/l) | **5.1** | **5.2** | **0.49 (0.14, 0.83)** | **5.2** | **5.2** | **0.42 (0.01, 0.84)** |
| 30-min glucose (mmol/l) | 8.3 | 8.6 | 0.24 (-0.13, 0.61) | **8.3** | **8.7** | **0.47 (0.11, 0.82)** |
| 120-min glucose (mmol/l) | 6.3 | 6.3 | -0.02 (-0.40, 0.35) | 6.3 | 6.8 | 0.23 (-0.25, 0.70) |
| OGTT AUC | 8.63 | 8.82 | 0.2 (-0.08, 0.48) | **8.62** | **9.09** | **0.38 (0.06, 0.69)** |
| Glucose intolerance (IFG+IGT) | 23 (22.5) | 11 (27.5) | ns | **16 (16.3)** | **16 (48.5)** | ***p*<0.001** |
| **Pregnancy (28 wks.)** | *n* =127 | *n* =44 |  | *n* =119 | *n* =39 |  |
| Height (cm) | 157 | 158.2 | 0.14 (-0.21, 0.48) | 157.7 | 156.3 | -0.27 (-0.63, 0.09) |
| Weight (kg) | **52.5** | **58.0** | **0.60 (0.26, 0.93)** | 53.5 | 53.7 | -0.03 (-0.44, 0.37) |
| BMI (kg/m<sup>2</sup>) | **21.3** | **23.3** | **0.52 (0.19, 0.86)** | 21.5 | 22.4 | 0.09 (-0.32, 0.50) |
| Gestational weight gain (GWG, kg) | 8.9 | 8.9 | 0.29 (-0.06, 0.65) | **8.5** | **10.1** | **0.42 (0.03, 0.81)** |
| Fasting glucose (mmol/l) | **4.3** | **4.9** | **1.63 (1.39, 1.87)** | **4.4** | **4.5** | **0.45 (0.04, 0.87)** |
| 60-min glucose (mmol/l) | **7.5** | **7.9** | **0.37 (0.02, 0.72)** | **7.1** | **9.4** | **1.53 (1.26, 1.81)** |
| 120-min glucose (mmol/l) | **6.2** | **6.6** | **0.42 (0.06, 0.78)** | **6.1** | **7.4** | **0.99 (0.67, 1.31)** |
| OGTT AUC | **7.6** | **8.26** | **0.66 (0.32, 0.10)** | **7.45** | **9.09** | **1.57 (1.38, 1.76)** |
| **24 Years (Post-Delivery)** | *n* =103 | *n* =33 |  | *n* =95 | *n* =25 |  |
| Height (cm) | 156.75 | 158 | 0.21 (-0.14 , 0.57) | 157.5 | 167.4 | -0.33 (-0.77, 0.12) |
| Weight (kg) | **49.9** | **60** | **0.70 (0.32, 1.10)** | 51.0 | 54.0 | 0.03 (-0.46, 0.52) |
| BMI (kg/m<sup>2</sup>) | **20.5** | **23.8** | **0.65 (0.23 , 1.07)** | 20.9 | 21.8 | 0.11 (-0.40, 0.62) |
| Overweight/obese *n* (%) | **17 (16.5)** | **15 (45.4)** | ***p*<0.01** | 25 (26.6) | 6 (24.0) | ns |
| Waist circumference (cm) | **72** | **78.7** | **0.76 (0.33 , 1.19)** | 75.7 | 78.3 | 0.21 (-0.14, 0.57) |
| Central obesity, >=80, *n* (%) | **21 (23.3)** | **18 (60)** | ***p*<0.01** | 28 (32.2) | 8 (33.3) | ns |
| Fasting glucose (mmol/l) | **4.9** | **5.3** | **0.77 (0.36 , 1.18)** | 5 | 5.3 | 0.42 (-0.07, 0.92) |
| 30-min glucose (mmol/l) | **7.9** | **8.8** | **0.66 (0.30 , 1.02)** | **8** | **8.7** | **0.56 (0.13, 0.99)** |
| 120-min glucose (mmol/l) | 5.7 | 6.3 | 0.36 (-0.06 , 0.78) | **5.7** | **6.2** | **0.42 (0.00, 0.85)** |
| OGTT AUC | **8.12** | **9.14** | **0.64 (0.29, 0.98)** | **8.12** | **9.04** | **0.57 (0.17, 0.97)** |
| Glucose intolerance (IFG+IGT) | **9 (8.7)** | **13 (38.2)** | ***p*<0.001** | 12 (12) | 7 (26.9) | ns |
| **F2 generation** | *n* =126 | *n* =43 |  | *n* =118 | *n* =38 |  |
| Male % * | 51.6 | 58.1 | - | 55.1 | 47.4 | - |
| Birthweight (g) | 2753 | 2780 | - | 2760 | 2805 | - |
| Birthweight Z-scores (INTERGROWTH) | -1.3 | -0.8 | 0.35 (-0.07, 0.77) | -1.5 | -1.4 | 0.11 (-0.26, 0.48) |

Data is presented as median and *n* (%) as appropriate along with mean Z difference (95% CI) by *t*-test. For proportions, significance is calculated by $\chi^2$ test, ns non-significant. The numbers (*n*) are different for different variables. The values for OGTT AUC have been divided by 100 for readability. Birthweight in g is
shown for context; the significance is shown for the sex and gestational age adjusted Z-scores (INTERGROWTH). Gestation weight gain was calculated as the difference between weight at 28wks and weight at 18-year follow-up.

### Pregnancy

Marriage and pregnancy were recorded by the field staff. At 28 weeks’ gestation, a fasting OGTT (75g anhydrous glucose, fasting, 60- and 120-mins venous samples) was performed. GDM was diagnosed by the IADPSG criteria (21) and appropriately treated. Neonatal anthropometry (F2) was performed using standardized methods.

### Post-delivery follow-up

At ∼24 years of age, F1 women underwent a 75gm OGTT, results were classified by ADA criteria (2). This activity was suspended due to the COVID pandemic (Mar 2020) and resumed in 2022. We used data collected until January 2024 for this analysis.

### Definitions

In F0 mothers, GDM was diagnosed by WHO 1985 criteria (75gm fasting OGTT) (22). In the F1 generation, GDM was diagnosed by IADPSG criteria (21). Given the small number of GDM cases (n=20/158, 12.6%), to increase statistical power for this paper, we defined ‘fasting hyperglycemia’ in F1 mothers as those in the highest quartile of fasting plasma glucose concentrations (FPG) (Q4 ≥ 4.7 mmol/l) and considered Q1+Q2+Q3 as ‘normoglycemia’. Similarly, we also calculated Q4 for the area under the curve (AUC) for OGTT glucose concentrations by the trapezoidal rule (Q4 ≥ 8.57 x 10^2^). We calculated similar Q4s for FPG (6-, 12-, and 18-years) and OGTT AUC (6- and 18-years).

### Statistical analysis

Our main analysis is to compare the life-course evolution of glycemia and body size measurements in women (F1) with and without pregnancy hyperglycemia at 28-wks gestation (fasting and OGTT AUC). Variables with skewed distributions were normalized by log transformation. All variables were converted to z-scores, and differences between normoglycemic and hyperglycemic participants were expressed in z-score units with 95% CIs and tested using *t*-test statistics. The life-course evolution of glycemic and size measurements is shown as serial line plots (mean ± SEM) for those with and without pregnancy hyperglycemia.

We calculated odds ratios to estimate the risk of having pregnancy hyperglycemia (FPG and AUC) for Q4 of FPG at 6, 12, and 18 years, and Q4 of AUC at 6 and 18 years. Similarly, we estimated the risk of being glucose intolerant (prediabetes+diabetes) at 24 years for Q4 of FPG at 6, 12 and18-years and 28 weeks gestation, and for Q4 of AUC at 6 and 18 years and 28wks gestation. The odds ratios are represented using forest plots.

All statistical analyses were performed using SPSS v26 and R 4.0.5 software.

### Ethics

The study was approved by village leaders and the KEM Hospital Research Centre Ethics Committee, Pune, India at all timepoints. Participants >18 years of age signed an informed consent. Children <18 years of age provided assent along with parental consent.

#### Data sharing statement

Data are available for additional analyses by applying to the corresponding author with a 200-word plan of analysis. Data sharing is subject to KEMHRC Ethics Committee approval and the Government of India’s Health Ministry Screening Committee (HMSC) permission.

## Results

### Cohort Description (figure 1, ESM figure S1)

F0 mothers (1993-96, n= 762) were 21.2 years old, 1.52 m tall, with a median BMI of 20.2 kg/m^2^ at 28wks gestation, and a third were primiparous. A 75g fasting OGTT was available on 598, median FPG was 3.9 mmol/l and 2h value was 4.3 mmol/l; only 3 were diagnosed GDM by WHO 1985 criteria. Three hundred and sixty-six live singleton daughters (birthweight 2.55 kg) and 396 sons (2.70 kg) were born in the study (F1, 1994-96). By February 2020, 195 F1 daughters had married, become pregnant and delivered. Our analysis includes 171 of these with fasting glucose and 158 with 75g OGTT data at 28 weeks gestation. They were 20.9 years old, 1.57 m tall, with a median BMI of 21.7 kg/m^2^, all were primiparous. Eighty-four live singleton daughters (2.78 kg) and eighty-five sons (2.75 kg) were born (F2). One hundred and thirty-six F1 daughters also attended a post-delivery follow-up (2019–24) including a 75g OGTT at an average age of 24.3 years, and a median BMI 21.3 kg/m^2^.

Compared to those not included in this analysis, the included F1 daughters had similar birthweight, and BMI and FPG at 6, 12, and 18y (ESM table 3).

### F1 daughters: glycemia and body size

At 28wks gestation, median FPG concentration was 4.4 mmol/l and 120 mins 6.3 mmol/l. Forty-four women fulfilled our definition of fasting hyperglycemia (Q4), and 39 using the AUC definition (Q4). Twenty women (12.6% of 158) were diagnosed GDM based on IADPSG criteria. At 24 years of age, 21 (15%) had glucose intolerance by ADA criteria (12 IFG, 5 IGT, 4 IFG+IGT).

### Glycemic and body size trajectories of F1 women with and without pregnancy hyperglycemia (Table 1, Figure 2)

At 28wks gestation, women with fasting pregnancy hyperglycemia were 20.3 years old, and were heavier (23.3 vs 21.2 kg/m2) compared to the normoglycemic women; height and gestational weight gain (8.9 vs 8.9 kg) were similar (Table 1). There was no difference in their mother’s glucose levels during pregnancy (F0) (Table 1). Women with pregnancy hyperglycemia had consistently higher levels of FPG and OGTT AUC starting from childhood (6 years), through puberty (12 years) and young adulthood (18 years) (Table 1, Figure 2). The prevalence of IFG was higher at 12 years (4.5% vs 0.8%) and 18 years (15% vs 4.9%), and they also had higher HbA1c concentrations at 15-17 years of age (mean diff +0.27 SD, p<0.07). Post-delivery, women with fasting pregnancy hyperglycemia continued to have higher glycemia, and a substantially higher proportion (38% vs 9%) had glucose intolerance (prediabetes) compared to those with pregnancy normoglycemia.

**Figure 2:**
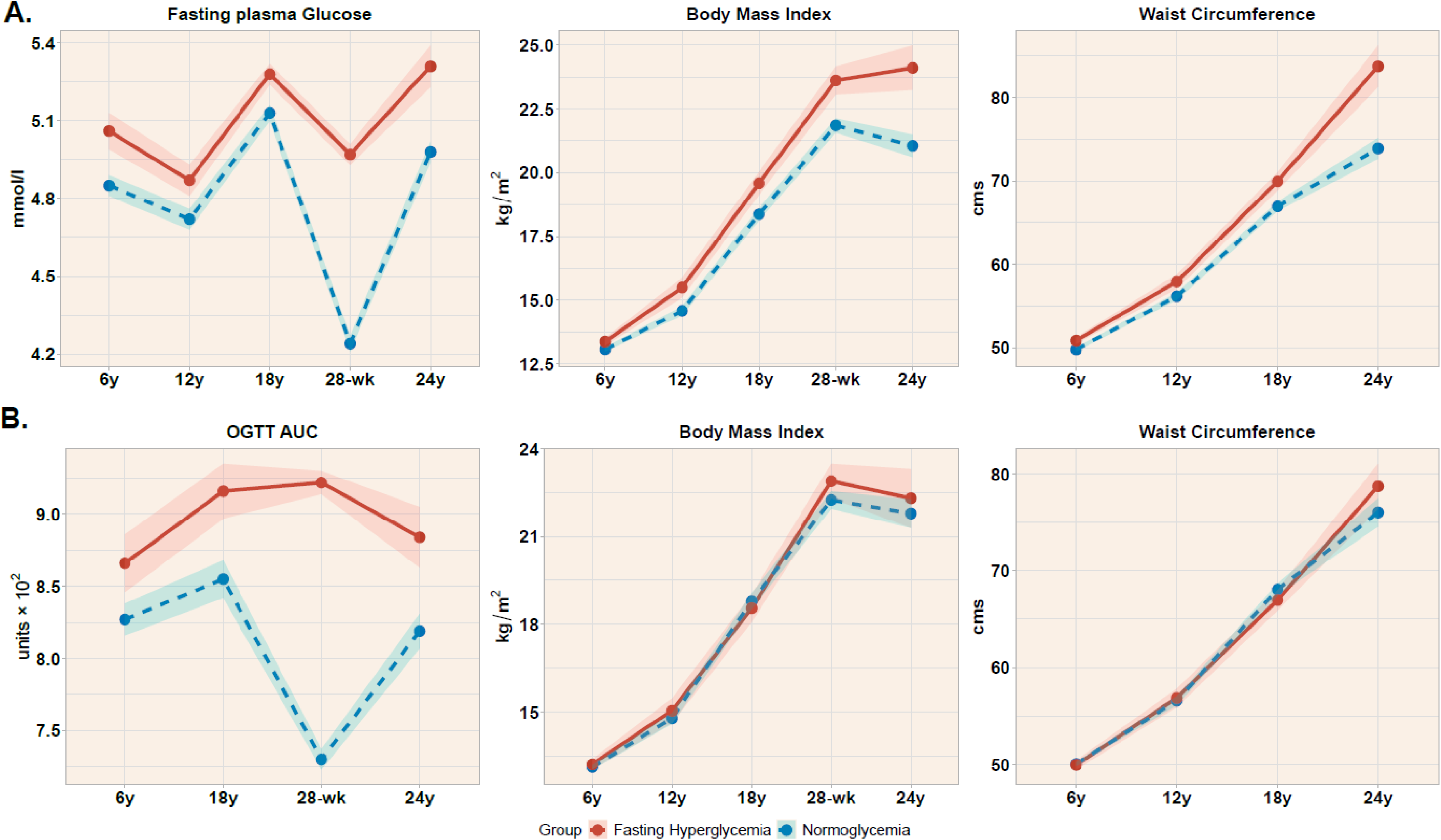
Life course tracking of glycemia and anthropometry in F1 daughters with and without pregnancy hyperglycemia. The figure shows serial mean ± SEM at different ages. Women are classified as hyperglycemic by Q4 of FPG (A), or Q4 of OGTT AUC (B) at 28wks gestation. Those with fasting pregnancy hyperglycemia had higher FPG, BMI, and waist circumference throughout. Those with OGTT AUC pregnancy hyperglycemia had higher glycemia throughout but not BMI and waist circumference.

Similarly, women with pregnancy OGTT AUC hyperglycemia had higher OGTT AUC at 6-years and 18-years, and higher HbA1c at 15-17-years. They continued to have higher glycemia post-delivery (table 1, figure 2).

### Prediction of pregnancy and post-pregnancy hyperglycemia by early life hyperglycemia (Figure 3 & 4)

Figure 3 shows that the prevalence of pregnancy hyperglycemia (both fasting and OGTT-AUC) progressively increased with increasing glycemia in early life; the forest plot shows that the cumulative risk was 2.22 [1.45-3.38] times higher for the Q4 of FPG and 2.88 [1.32-6.28] times for the Q4 of OGTT AUC. Figure 4 shows that the prevalence of post-pregnancy glucose intolerance progressively increased with increasing glycemia in pregnancy; the forest plot shows that the cumulative risk was 5.22 [2.40-11.33] time higher for the Q4 of FPG and 3.50 [1.36-8.97] times higher for the Q4 of OGTT AUC. All these risks remained similar and significant after adjusting for BMI or waist circumference.

**Figure 3:**
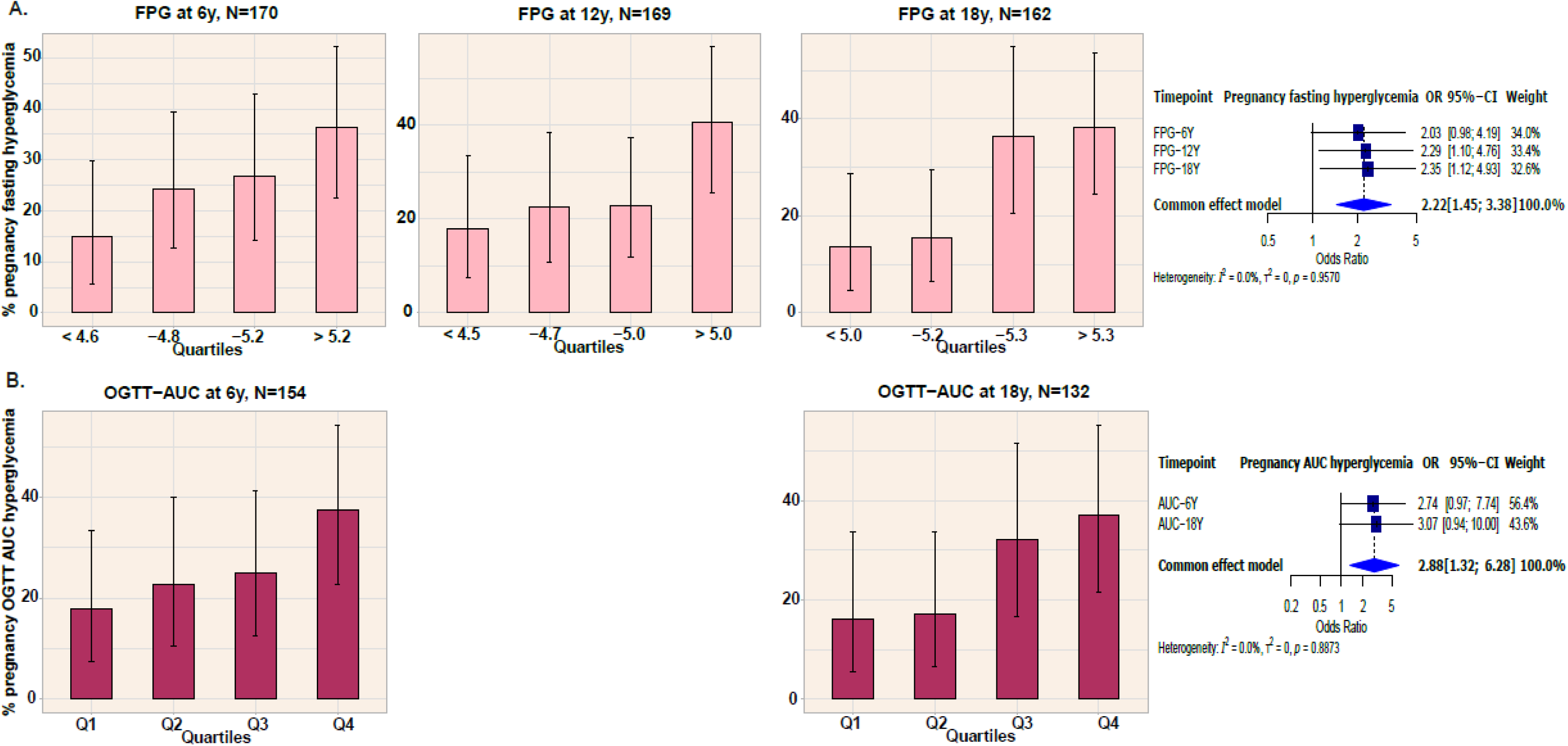
Prevalence of pregnancy hyperglycemia (28wks gestation) by quartiles of early life glycemia for fasting glucose (A) and OGTT AUC (B). The bars represent prevalence of pregnancy hyperglycemia with 95% CI. Please note OGTT AUC was not available for 12-year follow-up. There is a strong and graded association between early life glycemia and pregnancy glycemia. The forest plots show the ORs of pregnancy hyperglycemia for those in Q4 of early life glycemia

**Figure 4:**
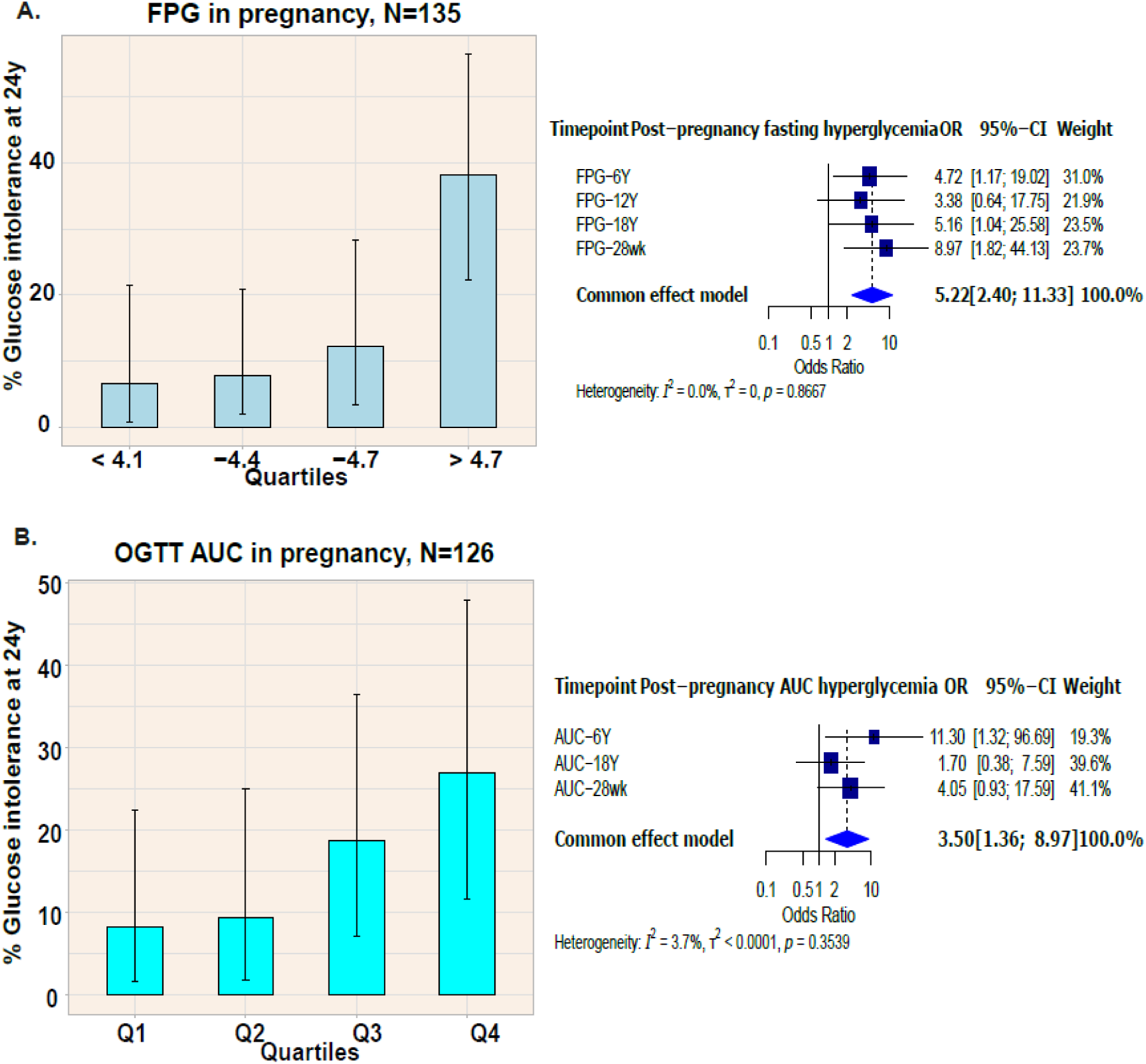
Prevalence of post pregnancy glucose intolerance (prediabetes, ADA criteria) by quartiles of pregnancy glycemia. The bars represent prevalence of post pregnancy glucose intolerance with 95% CI. There is a strong and graded association between pregnancy glycemia and post pregnancy glucose intolerance. The forest plots show the ORs of post-pregnancy glucose intolerance for those in Q4 of early life and pregnancy glycemia

### Trajectories of body size and growth (fig 2 and 5)

Our serial measurements from birth allowed us to study the life-course evolution of body size and growth in women with and without pregnancy hyperglycemia. Birth size was similar in those with and without fasting pregnancy hyperglycemia, however hyperglycemic women had experienced an early catch-up in height (at 2 and 6 years) and had higher weight, BMI and waist circumference from childhood through puberty, young adulthood, and post-pregnancy. It is noteworthy that, up to 18-years, the major issue in this population was ‘undernutrition’ (low BMI).

**Figure 5:**
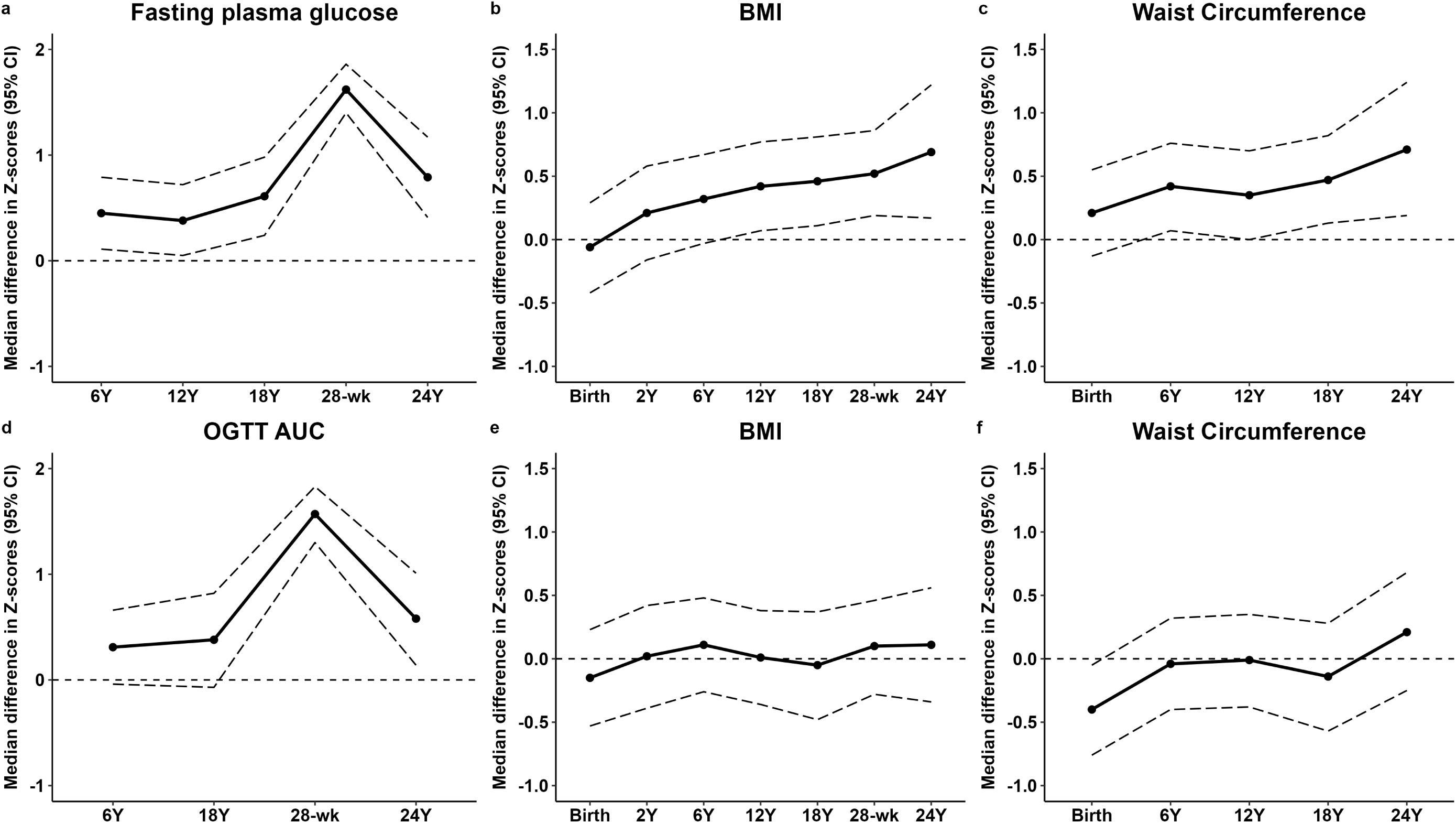
Differences between the two glycemic groups shown as plots of mean difference presented as Z-scores. The figure shows difference in fasting plasma glucose, BMI, and waist circumference in participants with (a-c) normoglycemia and fasting hyperglycemia, and (d-f) normoglycemia and OGTT AUC hyperglycemia. The points represent mean values with SEM as shaded region.

Women with pregnancy OGTT AUC hyperglycemia were lighter, shorter, and had smaller abdominal circumference at birth but their size after birth was no different than those with normoglycemia.

## Discussion

We present unique data to show that gestational hyperglycemia is only a window in the life-long hyperglycemic trajectory of that woman. We show tracking of fasting and post-glucose load glycemia (OGTT AUC) in female participants born in the PMNS birth cohort from their early childhood into puberty, early adulthood, pregnancy, and post-delivery. To the best of our knowledge, this is the first demonstration of such life course tracking. Our previous report on larger number of PMNS participants (male+female) showed that hyperglycemia at 18 years of age was significantly correlated also with higher maternal glycemia, which suggests that the tracking extends intergenerationally (20). These findings challenge the popular belief that ‘gestational’ hyperglycemia has a *de novo* onset during pregnancy and resolves after delivery. Thus, our findings have important implications for the definition, diagnosis, treatment, follow-up, and fetal programming effects of the condition called ‘GDM’.

The majority of reports in the literature concentrate on the increased risk of post-pregnancy diabetes in GDM women. Only a few studies have reported glucose metabolism in the pre-pregnancy period, but none in a life course model. Catalano’s classic studies of 38 women before and during early and late pregnancy showed that women subsequently diagnosed with GDM had higher glycemia and lower insulin sensitivity before pregnancy and maintained their differences from non-GDM women in early and late pregnancy (13). A Kaiser Permanente study in California showed that women diagnosed with GDM had higher BMI, higher glycemia, and dyslipidemia years before pregnancy, compared to women without GDM (14). Similarly, women diagnosed with GDM in the CARDIA cohort (15), MUSP cohort of aboriginal women in Australia (16), and the Cardiovascular Risk in Young Finns Study (17) had higher glycemia (glucose, HbA1c) before pregnancy. A recent demonstration of higher HbA1c values in early pregnancy in those subsequently diagnosed with GDM strongly suggests higher glycemia before pregnancy (23). Data from population-based administrative databases in Canada showed that women diagnosed with GDM during pregnancy already had elevated levels of fasting and random glucose, HbA1c, and dyslipidemia a few years before pregnancy (18,24). None of these studies have reported early childhood and intergenerational measurements. Outside pregnancy, many studies including Bogalusa Heart Study (25), the Early Bird study (26), i3C (27), and three Pune cohorts (20) have demonstrated tracking of glycemia from early life into young adulthood.

Ideas about the adverse clinical effects of gestational hyperglycemia have evolved since the 1960’s. The original concept was based on an increased post-partum risk of diabetes in the mother (28). Adverse pregnancy outcomes (3) and an increased risk of diabesity in the offspring were added later (29). Adverse pregnancy outcomes are mostly related to fetal overnutrition and consequent overgrowth (due to maternal glucose, lipids etc) and maternal co-morbidities (higher age, obesity, high blood pressure etc). The long-term risk of offspring diabesity is ascribed to fetal epigenetic programming (‘teratogenesis’) by the maternal metabolic milieu, in addition to the role of genetics (12). We now understand that the strongest window for epigenetic programming is pre- and periconceptional (preimplantation) (30, 31). Our data suggest that the ova and the early embryo of a woman subsequently diagnosed with gestational hyperglycemia will have been exposed to the yet undiagnosed hyperglycemia (and other metabolic problems) from an early age which could cause epigenetic modifications during gametogenesis, gamete maturation and early embryonic development and increase the risk of future diabesity. Thus, our findings have important implications for the timing of diagnosis and treatment of hyperglycemia to avoid programming of diabetes in the offspring and to curtail the escalating epidemic of diabetes in the young. The current practice of diagnosing and treating GDM in the third trimester partially reduces overgrowth of the baby but not the long-term risk of diabetes and obesity (10,11). Pre-gestational hyperglycemia in mice epigenetically reduced the expression of the TET3 gene (involved in DNA demethylation) and suppressed the expression of the *GCK* and other genes involved in insulin secretion in developing pancreatic islets, to increase the risk of diabetes in the offspring (32). Similar changes were observed in blastocysts of hyperglycemic women. The pancreatic islets from hyperglycemic donors showed epigenetic changes (DNA methylation) in the B-cells, which predisposed the recipient to future diabetes (33). Our preliminary investigation of circulating miRNAs in serial samples from women in the PMNS demonstrated a differential signature from early childhood in those with prediabetes at 18-years of age (34). These studies strongly support the thesis that the preceding lifelong dysglycemia in women diagnosed with ‘GDM’ will have already altered the ‘epigenetic profile’ in the ova and the embryo before diagnosis of GDM and increase the risk of diabetes in the offspring.

The main strength of our study is the serial metabolic and anthropometric data from childhood to pregnancy and the post-pregnancy period, spanning 30 years, collected using standardised methods and with high rates of follow-up. This allowed us to construct a trajectory that was not possible before. Our results show that the ‘glucostat’ is set very early in life for both fasting and post-glucose (OGTT) glycemia.

Limitations are that our study, being intense, was relatively small, and despite a moderately high prevalence of GDM (12.6%) there were only 20 cases. To increase statistical power, we defined hyperglycemia during pregnancy in our women using the upper quartiles of fasting glucose or the OGTT AUC. This could appear as a major limitation, but for the following reasons: 1) Very importantly, the HAPO study showed clearly that there are continuous and graded associations without any thresholds, between maternal glucose concentrations (fasting, 1h and 2h during an OGTT) and neonatal outcomes (macrosomia, hyperinsulinemia, hypoglycemia, and Caesarean section rates). Consensual and arbitrary statistical cut-points were defined in the absence of a relevant glycemic threshold for adverse outcomes which became the IADPSG criteria (21). Thus, both the exposures and outcomes in the HAPO study were centile-based, 2) Even after HAPO study and the IADPSG criteria, there is considerable disagreement about the criteria to diagnose GDM across the world. Though both WHO and ADA have accepted the IADPSG criteria, many countries have not (UK, Australasia, Denmark, Norway, and many centres in the USA and India), 3) Moreover, there was no representation in the HAPO study from the undernourished populations of the world. India, which is estimated to contribute a substantial number of GDM cases in the world (1) was not represented. Indian national average birth weight is 2.8 kg and intrauterine growth restriction is common, raising doubts about the applicability of the fetal overgrowth-based criteria from the high-income countries. The generalizability of our findings to other populations will need to be confirmed; however, available data (discussed above) indicates that the biology of life-course tracking is universal.

Our findings have the potential to change clinical thinking and public health practice. The preconceptional period provides an important opportunity to curtail developmental programming of diabetes, obesity and other non-communicable diseases (30, 31). This shifts the attention from the clinic to the community, from clinical practice to public health. Controlling lifestyle factors which promote diabesity during childhood and adolescence qualifies as ‘primordial’ prevention for the next generation. This will need cooperation between medical professionals, parents, schools, and governmental and non-governmental agencies to promote health literacy/education. This invites considerable efforts from the health system and expects a change in human behaviour. This is different from the currently popular practice of ‘primary’ prevention by targeting adults with prediabetes and obesity. Geoffrey Rose highlighted the larger benefits of intervening to favourably shift the curve rather than concentrating on the tail-end of the distribution (35). Fortification of flour with folic acid to prevent neural tube defects, and strict control of pre-gestational diabetes to prevent congenital anomalies are prime examples of successful primordial prevention.

In summary, we report novel data showing lifecourse tracking of glycemia from childhood in women diagnosed with gestational hyperglycemia. It is clear that gestational glycemia is only a window in the life-course evolution of metabolism. ‘GDM’ is not a *de novo* disorder of pregnancy as highlighted in a review many years ago (36). John Jarrett aptly commented that ‘gestational diabetes is simply a case of impaired glucose tolerance temporally associated with pregnancy’ (37). Maureen Harris also commented similarly (38). Our findings have implications for prevention of pre- and periconceptional epigenetic programming of diabetes and obesity which goes unattended in the current practice of diagnosing GDM in pregnancy (even at the booking visit) (39). A life-course ‘primordial’ approach to improve metabolic health of females from early life will be necessary to curtail the ‘vicious intergenerational cycle’ of diabesity in their offspring.

## Supporting information

This is the ESM file

## Data Availability

Data is available with Prof C S Yajnik for sharing to confirm our findings and for additional analyses by applying to the corresponding author with a 200-word plan of analysis. Data sharing is subject to KEMHRC Ethics Committee approval and Government of India Health Ministry advisory committee permission

## Acknowledgements

We thank the participants, collaborators, funding agencies, and the Diabetes Unit staff for their contribution towards this study. We would also like to thank Prof. Clive Osmond, MRC-LEU, University of Southampton, UK for his valuable insights into the statistical analysis.

## Authors’ contributions

CY and CF conceptualized the study. SB and RW were involved in statistical analysis. KJC, PY, RL contributed to conduct of the study. CY, CF, and RW prepared the manuscript. DB was involved in laboratory measurements. CY is the guarantor of this work and, as such, had full access to all the data in the study and takes responsibility for the integrity of the data and the accuracy of the data analysis. All authors approved the final version of the manuscript.

