## Supplementary material for "Diagnosing hyperglycemia (GDM) in pregnancy: closing the door after the horse has bolted?": This is the ESM file

**Electronic Supplemental Material**

**Index**

| **Sr No.** | **Item** | **Pg. No.** |
| --- | --- | --- |
| **1** | **ESM table 1:**  Methods for anthropometry and body composition measurements in children | 2 |
| **2** | **ESM table 2:**  Quality assessments for glucose measurements in the Pune Maternal Nutrition Study (1993- 2024) | 3 |
| **3** | **ESM table 3:**  Comparison between the women included in the analysis to those excluded | 4 |
| **4** | **ESM figure 1:**  STROBE flow diagram for the Pune Maternal Nutrition Study. | 5 |

**ESM table 1: Methods for anthropometry and body composition measurements in children**

| **Anthropometry** | | | |
| --- | --- | --- | --- |
| Measurement | Time point | Method | Least count |
| Weight | Within 72 hours of birth | Salter spring balance (Salter Abbey, Suffolk, U.K.) | Nearest 50 g |
|  | 6 monthly intervals up to 18-years, and 24-years | Electronic weighing scales (ATCO Healthcare Ltd, Mumbai, India) | Nearest 10 g |
| Crown heel length (Supine) | Within 72 hours of birth and 6 monthly upto 2 years | Portable Pedobaby Babymeter (ETS J.M.B., Brussels, Belgium) | Nearest 0.1 cm |
| Standing height | 6 monthly intervals from 2- 5 years | Portable Harpenden stadiometer (Microtoise, CMS Instruments Ltd, London, UK) | Nearest 0.1 cm |
|  | 6 monthly intervals from 5-18 years, and 24-years | Wall-mounted stadiometer (Microtoise, CMS Instruments Ltd, London, UK) | Nearest 0.1 cm |
| Waist Circumference | 6 monthly intervals from 5-18 years, and 24-years | Fiberglas tapes (CMS Instruments, London, U.K.). | Nearest 0.1 cm |
| Sum of skinfolds | Within 72 hours of birth | Harpenden skinfold calipers (CMS Instruments, London, UK)] | Nearest 0.2 mm |
|  | 6 monthly intervals up to 12-years, yearly upto 18-years, and 24-years | Harpenden skinfold calipers (CMS Instruments, London, UK)] | Nearest 0.2 mm |

**ESM table 2: Quality assessments for glucose measurements in the Pune Maternal Nutrition Study (1993- 2024).**

|  | **1994-97** | **2000-02** | **2006-08** | **2013-14** | **2019-2024** |
| --- | --- | --- | --- | --- | --- |
| **Glucose measurement** | | | | |  |
| Method | GOD-POD | GOD-POD | GOD-POD | GOD-POD | GOD-POD |
| Equipment | Spectrum; Abbott, Irving, TX | Spectrum; Abbott, Irving, TX | Alcyon; Abbott, Irving, TX | Hitachi 902, Roche Diagnostics GmbH, Germany | Hitachi 902, Roche Diagnostics GmbH, Germany |
| Centrifugation temperature | Room temperature | Room temperature | 4°C | 4°C | 4°C |
| Processed on | Same day | Same day | Same day | Same day | Same day |
| Internal QC | Yes | Yes | Yes | Yes | Yes |
| Internal CV (%) | <3 | <3 | <3 | <3 | <3 |
| External QC | NA | NA | BioRad EQAS | BioRad EQAS | BioRad EQAS |
| EQAS CV (%) | NA | NA | 3.4 | 2.7 | 2.7 |

GOD POD: Glucose oxidase peroxidase; QC: Quality control, CV: Coefficient of variation

**ESM table 3: Comparison between the women included in the analysis to those excluded**

|  | **Included (n=171)** | **Excluded (n=168)** | **p-value** |
| --- | --- | --- | --- |
| Gestation age (weeks) | 39.1 (38.1-40.0) | 39.1 (38.4-40.2) | 0.417 |
| Birthweight (kg) | 2.67 (2.48-2.90) | 2.64 (2.41-2.88) | 0.212 |
| Age at 6y (years) | 6.14 (6.04-6.23) | 6.14 (6.07-6.26) | 0.956 |
| BMI at 6y (years) | 13.0 (12.4-13.7) | 13.1 (12.6-13.7) | 0.391 |
| Fasting plasma glucose at 6y (mg/dl) | 87.5 (83.0-95.0) | 86.0 (81.0-93.0) | 0.071 |
| BMI at 12y (years) | 14.4 (13.2-15.8) | 14.4 (13.2-16.4) | 0.776 |
| Fasting plasma glucose at 12y (mg/dl) | 85.0 (81.0-90.5) | 85.0 (82.0-90) | 0.603 |
| BMI at 18y (years) | 18.0 (16.8-20.2) | 18.2 (16.5-20.4) | 0.911 |
| Fasting plasma glucose at 18y (mg/dl) | 93 (90-96) | 92 (89-95) | 0.100 |

Data is presented as median (25^th^-75^th^), p-values calculated using Mann-Whitney U test.

**ESM figure 1**: STROBE flow diagram for the Pune Maternal Nutrition Study (F1 female population).

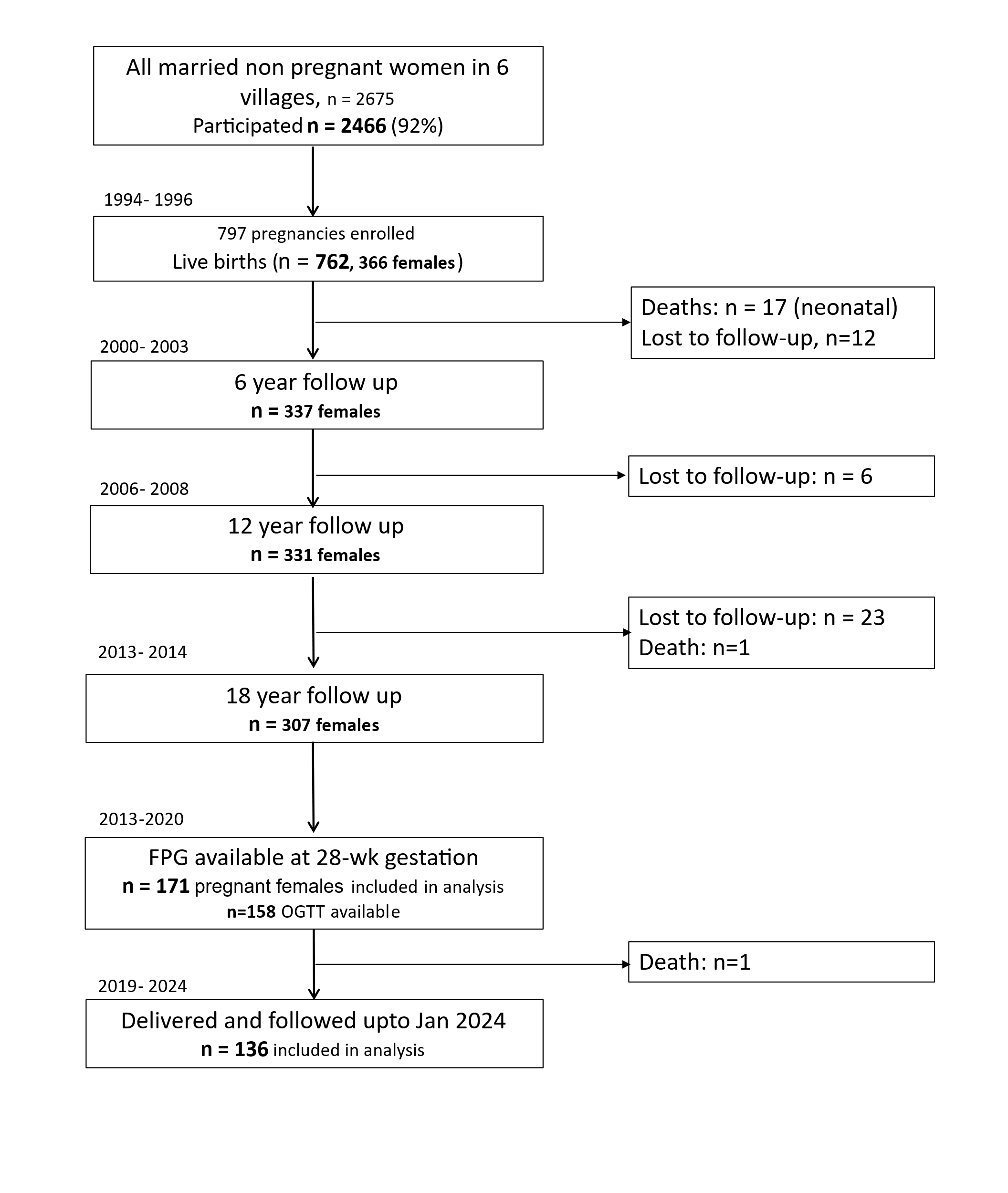
